# Effect of an integrated community-based intervention on antenatal care, incidence of malaria in pregnancy, adverse pregnancy and birth outcomes in rural Mali and Burkina Faso: The INTEGRATION cluster randomized trial

**DOI:** 10.64898/2026.03.19.26348775

**Authors:** Joel D. Bognini, Mahamadou Dembele, Biébo Bihoun, Kadiatou Koita, Sirima Traore, Toussaint Rouamba, Minh Huyen Ton Nu Nguyet, Oumou Coulibaly, Jean-Baptiste N’Tapke, Dario Scaramuzzi, Eve Worrall, Jenny Hill, Kassoum Kayentao, Halidou Tinto, Valérie Briand

## Abstract

**Background:** The WHO recommends at least eight antenatal care (ANC) contacts per pregnancy to improve maternal and child health. However, ANC coverage remains low in many settings in sub-Saharan Africa. Preventive measures, including intermittent preventive treatment of malaria with sulfadoxine-pyrimethamine during pregnancy (IPTp-SP), are crucial for reducing adverse pregnancy outcomes. This study assessed the effect of a community-based intervention on maternal and child health outcomes and malaria incidence in pregnancy in rural Mali and Burkina Faso.

**Methods:** This is a secondary analysis based on data from the INTEGRATION randomized cluster trial, encompassing 40 trial clusters (20 in Mali and 20 in Burkina Faso). The INTEGRATION intervention consisted of delivering IPTp-SP to pregnant women during seasonal malaria chemoprevention home visits from July to October over two consecutive years (2022 and 2023). Outside this four-month period, as in the control arm, IPTp-SP was given at the maternity clinic. Data were collected from the 40 study health facilities between January 2020 and June 2022 (pre-intervention period) and July 2022 and April 2024 (post-intervention period). The primary outcomes were ANC4+ coverage, cumulative incidence of malaria in pregnancy, and adverse pregnancy and birth outcomes, defined as any of the following: prematurity, low birth weight, stillbirth, or maternal death. A difference-in-differences model assessed the intervention’s effect on outcomes at the cluster level. Statistical significance was set at a 0.05 level.

**Results:** A total of 11,199 women in Burkina Faso and 35,351 women in Mali delivered during the study period. After adjusting for age and parity, the intervention did not show any statistically significant effect on ANC4+ coverage (Burkina Faso: -2.598; 95% CI [-13.400 – 8.202], Mali: -2.72; 95% CI [-14.35 – 8.91]), adverse pregnancy and birth outcomes (Burkina Faso: -0.194; 95% CI [-4.375 – 6.593], Mali: -0.36; 95% CI [-8.61 – 7.89]), or cumulative incidence of malaria in pregnancy (Burkina Faso: 125.56; 95% CI [-389 – 640], Mali: -12.68; 95% CI [-221 – 196]).

**Conclusion:** In both Burkina Faso and Mali, the four-month community-based intervention did not yield any statistically significant effect on maternal or pregnancy outcomes. Extending the duration of such interventions may be necessary to achieve meaningful reductions in the targeted outcomes.

**Trial registration:** Retrospectively registered on August 11th, 2022; registration # PACTR202208844472053 (Pan African Clinical Trials Registry: https://pactr.samrc.ac.za/Search.aspx). Protocol v4.0 dated September 04, 2023. Trail sponsor: University of Sciences Techniques and Technologies of Bamako (USTTB), Mali.

## Introduction

Since 2016, the World Health Organization (WHO) has recommended a minimum of eight contacts between pregnant women and qualified health personnel during their pregnancy, replacing the previous recommendation of four focused antenatal care (ANC) visits [1]. Antenatal care provides an opportunity to assess the woman’s health and the progress of her pregnancy, address her concerns, and advise her on recognizing potential pregnancy complications. It also facilitates the planning of childbirth with healthcare providers [1]. These recommendations are generally aimed at improving maternal and child health, in particular by reducing maternal and perinatal mortality [1].

In 2022, the neonatal mortality rate was 17 and 18 per 1,000 live births globally and in low-income countries, respectively. However, this rate was 26 per 1000 live births in the WHO African Region [2]. Preterm birth and intrauterine growth restriction, estimated at 10% both worldwide [3,4], are important factors contributing to neonatal mortality. They can be caused by maternal factors such as low weight gain, malaria, gestational hypertension and anaemia, which are regularly monitored and, where necessary, treated during pregnancy [5]. Also, preventive measures are recommended and delivered through ANC services such as intermittent preventive treatment with sulfadoxine-pyrimethamine during pregnancy (IPTp-SP). IPTp-SP is administered from the second trimester of pregnancy to prevent malaria, and its adverse effects on pregnancy and birth outcomes [1]; at least 3 doses during pregnancy are recommended. As IPTp-SP is primarily administered during ANC visits, the number of doses received by pregnant women is directly linked to the number of ANC visits [6]. Low ANC coverage and inadequate IPTp-SP use have been associated with increased risks of malaria in pregnancy, stillbirth, perinatal morbidity (including low birth weight and low APGAR score), postpartum haemorrhage, and maternal death [6–11]. In the WHO African Region in 2023, only 44% of pregnant women received three or more doses of IPTp-SP, which is below the WHO target of 80% [12].

In Burkina Faso and Mali, the 2016 WHO recommendations were adopted in 2018 and 2019, respectively, but not yet implemented at the national scale. In practice, at least four focused ANC visits are still widely used, with coverage remaining low (54 % in Burkina Faso and 30.5 % in Mali in 2022) [13,14]. These low ANC coverage rates are attributed to various barriers, including limited access to health facilities due to remoteness, poor road conditions, lack of transportation, and financial constraints [15–18]. To address these challenges and improve ANC and IPTp-SP uptake, with the ultimate goal of reducing pregnancy and child adverse outcomes, community-based interventions have been designed and evaluated. These interventions aim to encourage early ANC attendance (in the first trimester) and promote multiple ANC visits during pregnancy, facilitating the delivery of IPTp-SP [19–23]. The intervention delivered as part of the INTEGRATION (**IN**creasing the up**T**akE of IPTp-SP throu**G**h Seasonal Mala**R**i**A** Chemo-preven**TION** channel delivery - RIA2020S-3302, EDCTP funding) project is one of such strategies, consisting of community-based awareness about malaria prevention and antenatal care, and the administration of IPTp-SP in the community during seasonal malaria chemoprevention (SMC) rounds [24]. This project was designed as a randomized cluster implementation trial comparing the administration of IPTp-SP according to the standard of care at the maternity clinic *versus* in the community during four months of SMC in addition to ANC in rural Mali and Burkina Faso.

This study addresses one of the secondary objectives of the INTEGRATION project, aiming to evaluate the effect of the community-based intervention (SMC+ANC) on ANC coverage, adverse pregnancy and birth outcomes, and cumulative incidence of malaria during pregnancy.

## Methods

### Settings

The INTEGRATION project (RIA2020S-3302) was carried out in the Boussé (Burkina Faso) and Kangaba (Mali) health districts, located in areas where malaria transmission is seasonal. In 2023, in the Boussé health district, located in the Plateau Central region, 52.3% of women attended at least 4 ANC visits, 70.4% received three doses of IPTp-SP, and 55% experienced malaria during their pregnancy [20]. In 2022, in Kangaba health district, located in the Koulikoro region, 42.8% of women attended at least 4 ANC visits, 58.4% received three doses of IPTp-SP, and 60% experienced malaria during their pregnancy [13,14].

### Intervention

In each country, 20 health facilities (clusters) were randomly selected, with 10 assigned to the control arm and 10 to the intervention arm (see protocol [24] for more information). In the intervention arm, in addition to the routine activities carried out by health care workers at the facility (i.e., pregnancy and ANC awareness and counselling, as well as delivery of usual preventive and curative measures), a community-based intervention was implemented. It consisted of delivering IPTp-SP using the SMC delivery channel. This platform was also used to provide advice to women on the importance of antenatal care and refer them to these services if necessary. SMC consists of door-to-door visits during which children aged 3-59 months receive antimalarial drugs (amodiaquine + sulfadoxine-pyrimethamine) for the prevention of malaria four times during the high transmission period (from July to October). As part of the INTEGRATION trial, pregnant women were identified during SMC home visits. Those who had already initiated ANC and received a dose of IPTp-SP at least 28 days earlier were offered to receive IPTp-SP at home. Pregnant women who had not yet started ANC, or received a dose of IPTp-SP, or presented with suspected symptoms were referred to the health facility for ANC initiation, or the first dose of SP, or curative care. All identified pregnant women received counselling on malaria prevention and the importance of ANC. Outside of the 4 months of SMC, women in the intervention arm received IPTp-SP at the maternity ward, according to the routine standard of care.

In the control arm, no activities were conducted by the INTEGRATION team in addition to standard ANC practices at the health facility. Routine activities consisted of health centre-based ANC provided by health care workers, along with pregnancy and ANC awareness and counselling, and delivery of IPTp-SP.

The INTEGRATION project was conducted for two consecutive years (2022 and 2023), with the community-based intervention delivered during SMC visits for four consecutive months per year (from July to October).

#### Conceptual framework

Our main hypothesis was that the community-based intervention (SMC+ANC combined strategy) would increase IPTp-SP coverage; by receiving more doses of IPTp-SP, women would be better protected against malaria and its adverse effects on pregnancy and childbirth. About the expected effect of the intervention on ANC coverage, two hypotheses were put forward: on one hand, raising women’s awareness on the importance of antenatal care and referring them to a health facility when they had not yet received the first dose of IPTp-SP or had not yet attended their first ANC visit could increase ANC coverage; on the other hand, the administration of IPTp-SP to pregnant women in the community could discourage them from seeking antenatal care (as they would no longer need to travel to a health facility to receive IPTp-SP), leading to a decrease in ANC coverage. In the end, assuming improved coverage of both IPTp-SP and antenatal care, women could benefit from better monitoring by health care workers and improved access to assisted deliveries in health facilities, thereby reducing adverse pregnancy outcomes (e.g., low birth weight, prematurity, maternal mortality). These different hypotheses are represented in the conceptual framework below (Fig 1).

**Fig 1.**
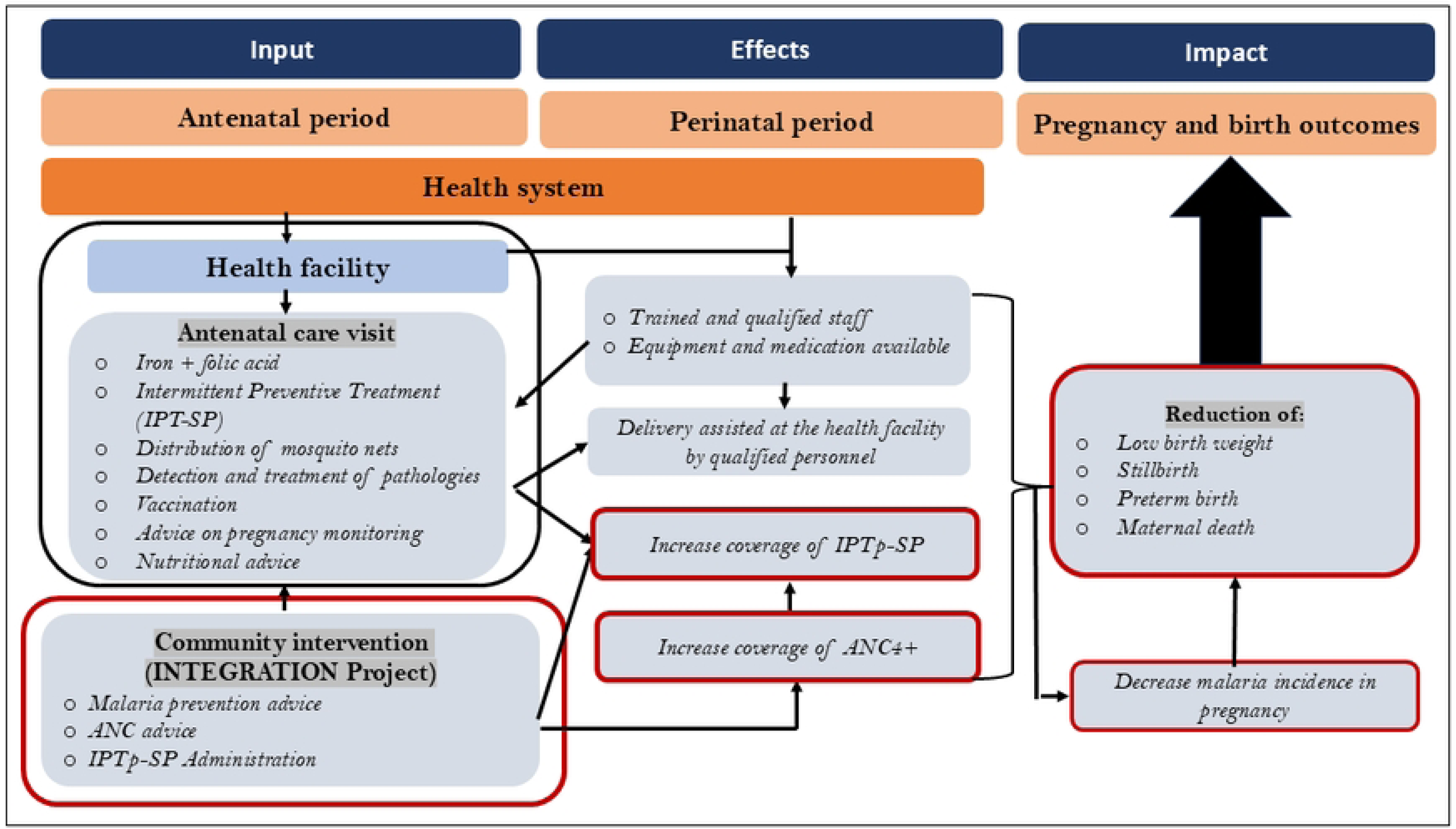
Conceptual framework: Assumptions and pathways regarding the effect of the community intervention on antenatal care, malaria during pregnancy, and pregnancy and birth outcomes.

### Data sources

In each country, routine data were collected retrospectively from January 2020 to June 2022, and prospectively from July 2022 to April 2024 in the 20 study health facilities in each country. Routine data were assessed from 1^st^ August 2022 to 10 April 2024. Consequently, the year 2022 includes both the pre- and post-intervention periods. The data were individual and were extracted from all ANC and delivery registers by the INTEGRATION project staff. The following data were extracted from the ANC registers: date of visit, actual number of ANC visits, pregnant woman’s marital status, age, occupation, and education level. The following data were extracted from the delivery registers: date of delivery, history of pregnancy (gravidity, parity, and still birth), birth weight, and occurrence of still birth, preterm birth, and maternal death. No link could be established between the two data sources. While the number of malaria cases in pregnancy was extracted from the ANC registers in Mali, we used aggregated data from District Health Information Software version 2 (DHIS2) to document the number of malaria cases in Burkina Faso.

#### Study outcomes

Three main outcome variables were defined for this analysis: ANC coverage, cumulative incidence of malaria in pregnancy, and adverse pregnancy and birth outcomes.

ANC4+ coverage was defined as the proportion of women who attended at least four antenatal care visits. It was calculated as the number of women with at least four ANC visits divided by the total number of deliveries in each health facility. For each country, it was aggregated to obtain: 1) annual estimates, and 2) overall estimates by arm and period of intervention, leading to an estimate of 20 coverage rates (10 in each study arm) before the intervention (January 2020 to June 2022) and 20 coverage rates (10 in each study arm) after the intervention (July 2022 to April 2024).

The proportion of adverse pregnancy and birth outcomes, namely maternal death, preterm birth, stillbirth, and low birth weight were combined and treated as a composite variable; the woman was considered as having an adverse pregnancy/birth outcome if at least one of the above-mentioned outcomes was reported. We selected maternal and perinatal mortality, prematurity, and low birth weight because they often share common causes (such as malaria, hypertension, etc.) that can be prevented or managed during antenatal care. Combining these events also makes it easier, for statistical power reasons, to evaluate the effect of an intervention on relatively rare events. The denominator was the number of women who delivered at the study health facilities. Annual estimates, as well as overall estimates by arm and intervention periods, were obtained.

The cumulative incidence of malaria in pregnancy was calculated as the number of malaria cases reported in pregnant women during each study period (i.e., pre- and post-intervention periods) divided by the total number of women who attended their first ANC visit during the same period. Annual estimates, as well as overall estimates by arm and period of intervention, were obtained.

#### Independent variables

The following sociodemographic characteristics and history of previous pregnancies were used: woman’s age (categorized according to the quartile into four groups: < 21 years, 21-25 years, 26-30 years, and > 30 years), parity (categorized as primiparous, secundiparous, or multiparous (three or more pregnancies), history of stillbirth (yes or no). The following variables were coded as follows: time: 0 = pre-intervention, 1 = post-intervention, and group: 0 = control arm, 1 = intervention arm. The pregnant woman’s education level, occupation, and marital status could not be used as they contained a high number of missing data.

### Statistical analyses

Analyses were performed using Stata 15.1. First, we described the characteristics of women who delivered in each health district. Then, annual trends in ANC4+, pregnancy and birth adverse outcomes, and cumulative incidence of malaria in pregnancy were calculated from 2020 to 2023. The data collected in 2024 is not presented here as it only covers four months of the year (January–April). Finally, a difference-in-differences (DiD) model was used to assess the effect of the INTEGRATION intervention on ANC4+ coverage, adverse pregnancy and birth outcomes, and cumulative incidence of malaria in pregnancy from 2020 to 2024. This type of model allows any difference between the trial arms (i.e., control *vs.* intervention) to be accounted for before the intervention implementation. The estimation of the effect of the intervention is based on the DiD coefficient (the difference between the estimated counterfactual value of the indicator in the intervention arm and the observed value of the indicator in the intervention arm). The formula for the difference-in-differences model is presented below [25]:

Y*=*β*0 +* β*1*Time +* β*2\**Intervention + β*3*\*Time*intervention β*4*\*Covariates *+* ɛ

Where Y is the outcome, β0 is the baseline average, β1 is the time trend in the control arm, β*2* is the difference between the two arms in the pre-intervention period, β*3* is the coefficient of DID estimating the interaction between time and intervention in the intervention arm, β*4* is the trend covariates, and *ɛ* is the error term. A positive DiD coefficient indicates an increase in Y in the intervention arm compared to the control arm, while a negative coefficient indicates a decrease. A significance level of 0.05 was set for statistical analyses.

#### Ethics

The study protocol was approved by the Institutional Review Boards/Ethics Committees and local health authorities in both countries (No 2022/83/CE/USTTB for Mali, No 2022/03-051/MSHP/MESRI/CERS for Burkina Faso) and by the Liverpool School of Tropical Medicine (22-006). Informed consent has been obtained from the participants, their parents, and legally authorized representatives in this study. Only the participant’s anonymous study ID was captured in the database. The database used in this study is available at the University of Mali, and access can be obtained after requesting authorization from the principal investigator.

## Results

A total of 11,199 pregnant women who delivered at the study health facilities in Burkina Faso, and 35,551 in Mali were included in this study. In Burkina Faso, the mean age of women was 26 (SD: 6) years, while in Mali it was 24 (SD: 5) years across both intervention and control arms. In Burkina Faso, in the intervention arm, most of the women (42.7%) were primiparous, with a previous abortion rate of 3.9%. In the control arm, 40.2 % were primiparous and 1.4% had experienced an abortion. In Mali, over 75% were multiparous and about 2% had experienced a previous infant death in both arms (Table 1).

**Table 1.** Characteristics of the study participants of Burkina Faso and Mali.

| Characteristics | Burkina Faso |  | Mali |  |
| --- | --- | --- | --- | --- |
|  | Intervention<br>n (%) | Control<br>n (%) | Intervention<br>n (%) | Control<br>n (%) |
| **Mean age (SD)** | 26.26 (6.37) | 26.62<br>(6.64) | 24.30 (5.93) | 24.10 (5.64) |
| **Age of the mother**<br>**(years)** |  |  |  |  |
| < 21 | 1264 (22,74) | 1252<br>(22,72) | 6249 (34.54) | 5913 (35.40) |
| 21-25 | 1534 (27,59) | 1447<br>(26,26) | 5073 (28.04) | 4697 (28.12) |
| 26-30 | 1431 (25,74) | 1359<br>(24,66) | 4389 (24.16) | 4282 (25.63) |
| >30 | 1330 (23,93) | 1453<br>(26,37) | 2381 (13.16) | 1812 (10.85) |
| **Parity** |  |  |  |  |
| Primiparous | 2371 (42.72) | 2209<br>(40.22) | 955 (5.20) | 448 (2.88) |
| Secundiparous | 893 (16.09) | 814 (14.82) | 3365 (18.31) | 3673 (21.64) |
| Multiparous (≥3) | 2286 (41.19) | 2496<br>(44.96) | 14061<br>(76.50) | 12809<br>(75.48) |
| **Previous**<br>**pregnancy/child** |  |  |  |  |
| Live birth | 612 (92.17) | 1064<br>(95.43) | 15489<br>(96.43) | 13985<br>(97.36) |
| Abortion | 26 (3.92) | 16 (1.43) | 139 (0.87) | 83 (0.58) |
| stillbirth | 11 (1.66) | 8 (0.72) | 102 (0.63) | 48 (0.33) |
| died | 15 (2.26) | 27 (2.42) | 333 (2.07) | 248 (1.73) |

**Table 2.**
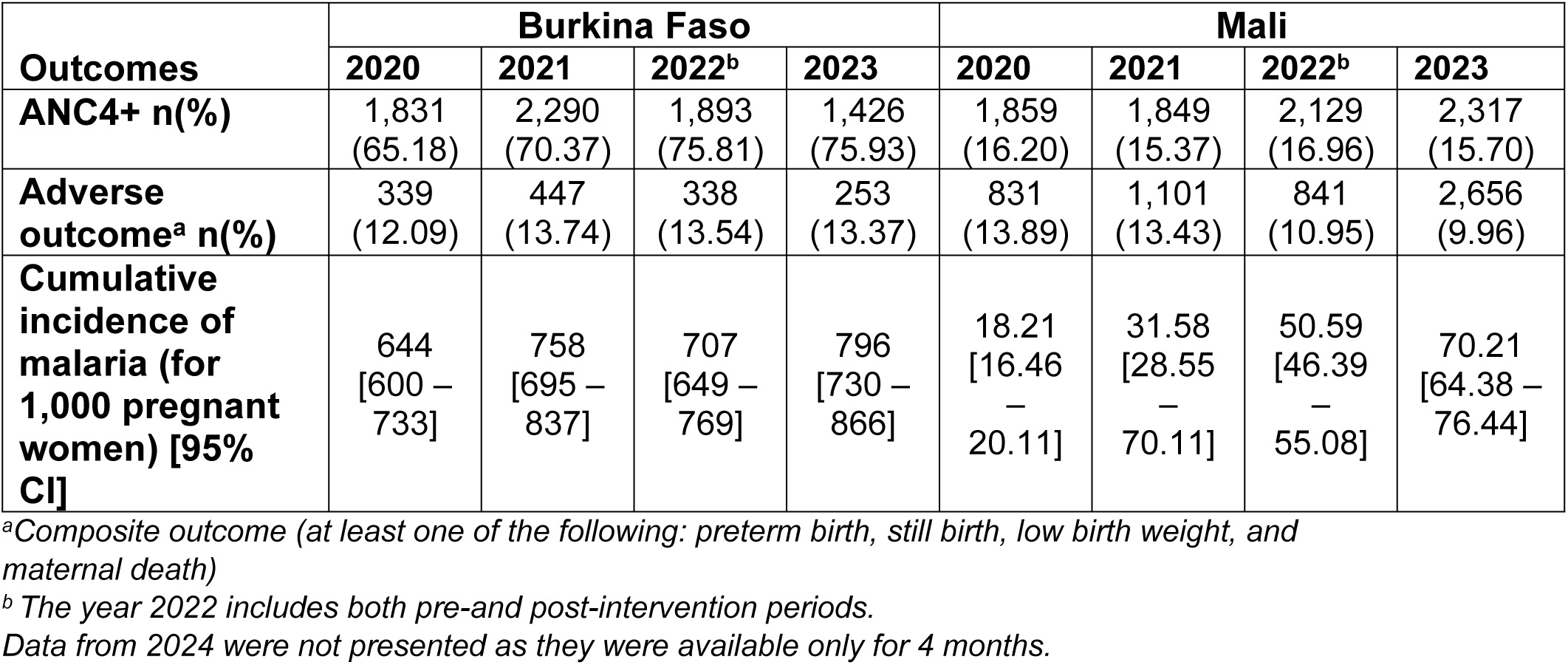
Trends in ANC4+ coverage, adverse pregnancy and birth outcomes, and cumulative incidence of malaria in pregnancy from 2020 to 2023 in Burkina Faso and Mali.

Between 2020 and 2023, the Boussé health district (Burkina Faso) recorded an increase in ANC4+ coverage, rising from 65% to 76%. Also, adverse pregnancy and birth outcomes and cumulative incidence of malaria among pregnant women increased from 12.1% to 13.4% and from 644 to 796 malaria cases per 1,000 pregnant women at risk, respectively. In Kangaba health district (Mali), the ANC4+ coverage slightly decreased from 16.2% in 2020 to 15.7% in 2023. Also, the rate of adverse pregnancy and birth outcomes decreased from 13.4% to 10%. However, the cumulative incidence of malaria among pregnant women increased from 18.2 malaria cases per 1,000 pregnant women to 70.2 per 1000 pregnant women at risk (Table 2). The evolution over time of the various indicators related to pregnancy and children (i.e., maternal mortality, premature births, stillbirths, and low birth weight) is presented for each indicator individually in Supplementary Table 1.

The INTEGRATION intervention did not show a statistically significant effect on any of the outcomes studied in Burkina Faso. While there were slight decreases in ANC4+ coverage (-2.6, 95 % CI [-13.2; 8.5]) and adverse pregnancy and birth outcomes (-0.19, 95 % CI [-4.4; 6.6]) in the intervention arm compared to the control arm, these were not statistically significant after adjusting for women’s age and parity. Malaria incidence increased by 125.6 (95 CI % [-389; 640]) in the intervention arm compared to the control arm, although this was also not statistically significant. In Mali, there was no statistically significant effect of the intervention on any of the studied outcomes. While the ANC4+ coverage and adverse pregnancy and birth outcomes decreased in the intervention arm compared to the control arm by 2.7 (95 % CI [-13.2; 8.5]) and 0.19 (95 % CI [-4.37; 6.59]), respectively, neither change was statistically significant after adjusting for women’s age and parity. Malaria incidence in pregnancy also decreased by 12.8 (95 % CI [-221; 196]) in the intervention arm compared to the control arm, but it was not statistically significant (Table 3). Illustrations of the intervention’s effect are presented in Supplementary figures (**Supplementary Fig. 1.** Illustration of the effect of the INTEGRATION intervention on ANC4+ coverage in Burkina Faso and Mali, **Supplementary Fig. 2**. Illustration of the effect of the INTEGRATION intervention on pregnancy/birth adverse outcome in Burkina Faso and Mali, and **Supplementary Fig. 3**. Illustration of the effect of the INTEGRATION intervention on the cumulative incidence of malaria in pregnancy in Burkina Faso and Mali).

**Table 3.**
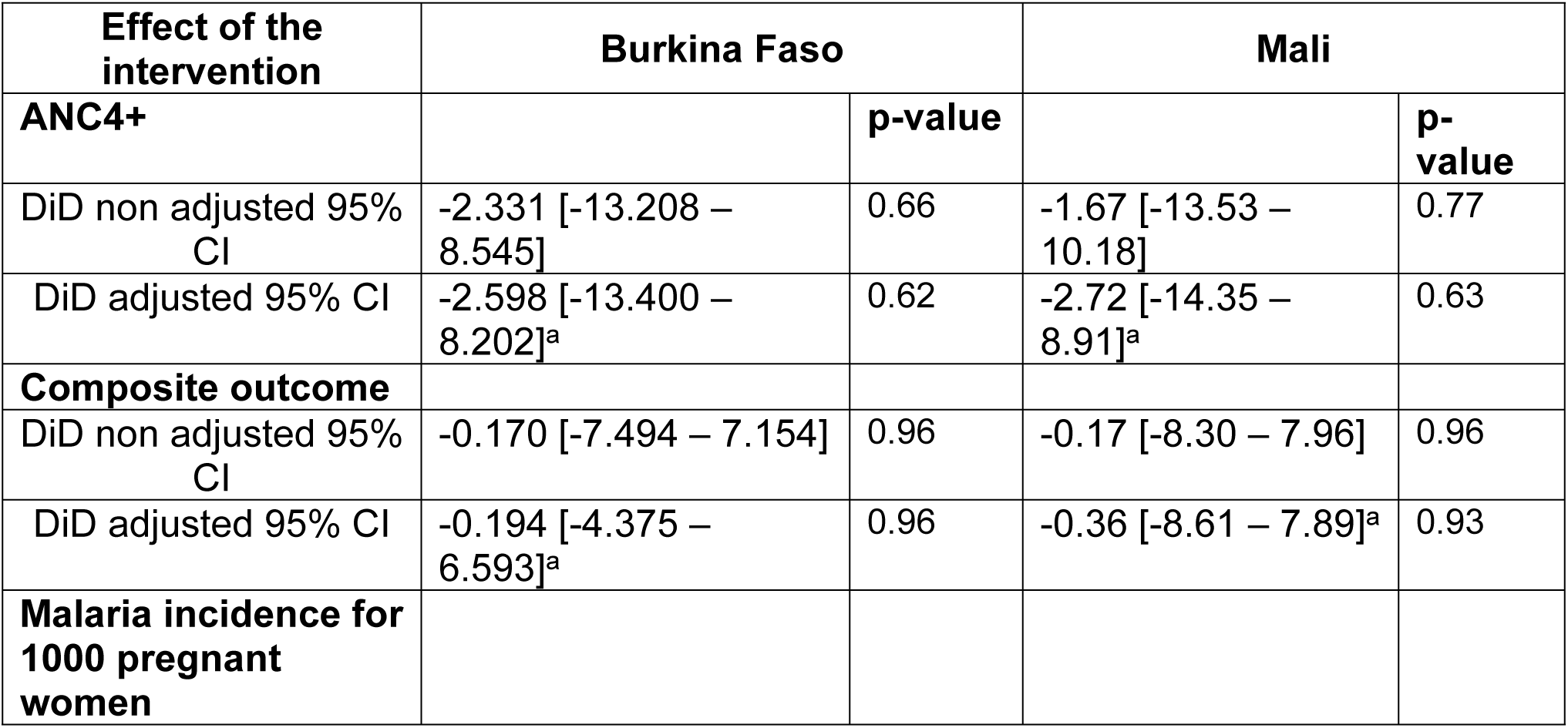
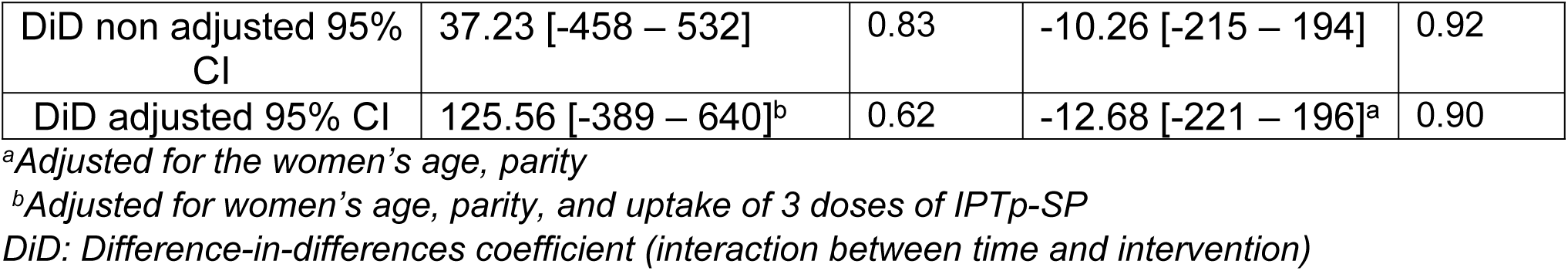
Effect of the INTEGRATION intervention on ANC4+ coverage, adverse pregnancy and birth outcomes, and cumulative incidence of malaria in pregnancy.

## Discussion

The study didn’t show any effect of the INTEGRATION intervention - integrating SMC and ANC for delivering IPTp-SP in the community on ANC4+ coverage, adverse pregnancy and birth outcomes, and cumulative incidence of malaria among pregnant women in Burkina Faso and Mali.

In this study, we hypothesized that the intervention would impact ANC4+ coverage (positively or negatively). In the Boussé health district (Burkina Faso), ANC4+ coverage showed a slight increase between 2020 and 2023. However, the observed trend was similar in both the intervention and control arms based on the DiD analysis. Given the high baseline ANC4+ coverage, demonstrating an effect of the intervention would have required a substantial increase in ANC4+ in the intervention arm compared to the control arm, which was not observed. A combination of health system factors may have influenced this observed increase in ANC4+ coverage. Indeed, since 2016, Burkina Faso has implemented several initiatives to improve maternal and child health, including free healthcare for pregnant women, strengthening of health facility with increased ratio of the staffing (a midwife per inhabitants increased from 4,831 in 2019 to 4,604 in 2023; a nurse per inhabitants increased from 3,074 to 3,014 in 2023), and improved access to care (average radius of action of health facility decreased from 4.1km to 3.9km) [13]. Additionally, the number of community health workers (CHWs) increased from 17,648 in 2019 to 18,721 in 2023, facilitating community-based interventions, including awareness-raising campaigns on the benefits and importance of ANC [26]. These nationwide efforts have likely contributed to the observed increase in ANC4+ coverage. Given that the INTEGRATION intervention was implemented in the community over a relatively short period of four months, it may not have had a sufficiently long duration to exert a significant effect on ANC4+ coverage in the intervention arm compared to the control arm.

In the Kangaba health district (Mali), ANC4+ coverage remained relatively stable between 2020 and 2023, and at a very low level. This stability may be attributed to several factors. Over this period, healthcare resources saw only a modest increase. The density of midwives and obstetric nurses improved slightly between 2020 and 2022, with the ratio rising from one health worker per 1,532 inhabitants to one per 1,498 [14,27]. Furthermore, only 0.7 % of the population experienced a reduction in distance to the nearest health facility from more than 5 km to less than 5 km [14,27]. As for Burkina Faso, we did not evidence any effect (either positive or negative) of the INTEGRATION intervention on ANC4+ coverage, as shown by the non-significant DiD coefficient.

It is important to note that, according to the WHO’s 2016 ANC recommendations, home visits conducted by CHWs for pregnant women can be considered ANC visits. In this study, home visits during which pregnant women received IPTp-SP, health education, and support, were not explicitly included in the calculation of ANC4+ coverage, as the WHO recommendations had not yet been implemented. Had these visits been counted, we might have observed an increase in ANC4+ coverage attributable to the intervention. Our results in Burkina Faso and Mali differ from those observed in a Kenyan study that reported a 26% statistically significant increase in ANC utilization following a community intervention involving CHWs. The intervention in Kenya consisted of raising awareness among pregnant women in the community to attend four ANC visits and carry out four tests (syphilis, haemoglobin level, malaria, and HIV) by four months during their pregnancy [28]. In another study conducted in Ethiopia, a 16% increase in ANC utilization was observed following a women’s group intervention [29]. The intervention in Ethiopia aimed to sensitize pregnant women within their communities with key messages, including safe childbirth practices and the importance of utilizing maternal health services. These differences in effect between our study and those cited above could partially be explained by the shorter duration of our intervention (four months per year over two years), compared to the twelve-month intervention in Kenya and the seventeen-month intervention in Ethiopia.

In Burkina Faso, a slight increase in the proportion of adverse pregnancy/birth outcomes was observed between 2020 and 2023, while a slight decrease was reported in Mali during the same period. We hypothesized that raising women’s awareness of the importance of antenatal care and referring them to a health facility when they had not yet received the first dose of IPTp-SP could improve both IPTp-SP and ANC coverage. This, in turn, could enable better pregnancy monitoring by health workers and ultimately increase the number of facility-based deliveries, as well as better prevention of malaria, thereby reducing adverse pregnancy and birth outcomes. However, we did not evidence such a reduction in adverse pregnancy/birth outcomes in either Burkina Faso or Mali. Our results contrast with those of a study conducted in Ghana that showed an improvement in health equity for pregnant women living in rural settings by bringing healthcare services directly to communities. That study’s success stemmed from reducing barriers to access (cost, time, socio-cultural) and promoting practices like antenatal care, good nutrition, hygiene, and facility-based delivery, leading to better pregnancy outcomes (fewer still births and miscarriages reported in this study) [30]. Again, these differences in findings may be attributable to the duration of the community intervention in Burkina Faso and Mali, which was relatively short, compared to the five-year duration of the intervention in the Ghanaian study. These results may also be explained by a different definition of the indicator of interest between the two studies, with our indicator including a greater number of adverse events, but possibly also fewer events specifically related to the care provided during antenatal visits.

In the present study, the cumulative incidence of malaria in pregnancy was observed to be far higher in the Boussé district in Burkina Faso compared to the Kangaba district in Mali. However, in both countries, malaria incidence evolved similarly over time in both study arms, with an increase observed in Burkina Faso and a decrease in Mali. This finding differs from the results of a meta-analysis including studies on malaria in pregnancy conducted between 2020 and 2023 in sub-Saharan Africa, Latin America, and Asia, reporting that the prevalence of malaria during pregnancy remained relatively stable [31]. Also, the INTEGRATION intervention did not demonstrate a statistically significant effect on the cumulative incidence of malaria among pregnant women in either country.

We acknowledge some limitations to our study. First, the use of several data sources across the two countries may have introduced bias and affected cross-country comparisons, particularly regarding malaria incidence. However, this should not have influenced the within-country before/after intervention comparisons. Second, we did not collect information on factors that could influence the three indicators of interest—such as the quality of ANC services, the quality of institutional delivery care, or women’s socioeconomic status. The absence of these data may have introduced bias, particularly if these factors differed between the pre- and post-intervention periods. However, to our knowledge, no other large-scale intervention on malaria or maternal health took place in the two districts during the study period. Third, the quality of the data extracted from the antenatal care and delivery registries was suboptimal in both countries. Indeed, several variables contained missing values, which may have introduced bias into the indicator estimates. Nevertheless, the large sample size likely helped to reduce this bias. In addition, we did not compare trends in deliveries— which served as denominators— over time, either before or after the start of the intervention, between the two study arms studied in each country. Also, marked differences in the total number of deliveries across health facilities in each arm could have resulted in inaccurate estimates of ANC4+ rates and adverse pregnancy and birth outcomes. Finally, the limited number of individual adverse pregnancy outcomes precluded a direct comparison between study arms; consequently, a composite variable was employed to evaluate the intervention’s effect.

## Conclusion

Our study revealed an overall increase in ANC4+ coverage in Burkina Faso, while in Mali, ANC4+ coverage remained relatively stable and low. We did not evidence any significant effect of the four-month community-based intervention on ANC4+ coverage, adverse pregnancy and birth outcomes, and cumulative incidence of malaria among pregnant women in either country. However, community-based interventions serving as platforms to promote ANC, particularly when implemented over a longer period (such as a full year), may offer significant benefits to women. This is especially relevant in light of the new WHO recommendations that emphasize the importance of every interaction with pregnant women, even those occurring outside health facilities.

## Data Availability

The database used in this study is available at the University of Mali, and access can be obtained after requesting authorization from the principal investigator.

## Acknowledgements

The authors wish to thank the Ministry of Health of Mali and Burkina Faso for giving us access to DHIS2 data, the health district of Kangaba (Mali) and Boussé (Burkina Faso) for their help in understanding the local context, and the INTEGRATION project for collecting routine data.

## Authors’ contributions

J.D.B. and V.B. conceptualized and designed the study. R.T and M.Y. supported the methodology. J.D.B. analysed and interpreted data, and wrote the first draft of the manuscript.

J.N., B.B., M.D., K.Ko, O.C., S.T, J.H., E.W., D.S., K.K., H.T., and V.B substantially revised the content of the manuscript.

All authors approved the submitted version and agreed both to be personally accountable for their own contributions and to ensure the accuracy or integrity of any part of this work.

